# Psychiatric partner resemblance and its socioeconomic component across the life course

**DOI:** 10.64898/2026.09.11.26362836

**Authors:** Saeid Rasekhi Dehkordi, Martin Dalgaard Villumsen, Dorte Helenius, Wouter Peyrot, Thomas Werge, Alfonso Buil, Abdel Abdellaoui

## Abstract

Partner resemblance influences and reflects the social, genetic and health structure of human populations. Using Danish registers on 787,658 couples, we quantified partner resemblance for ten psychiatric disorders and socioeconomic status (SES), measured up to a year before first childbirth and across the life course. Psychiatric partner correlations ranged from 0.08 for eating disorders to 0.36 for schizophrenia, below those for education (0.63) and income (0.57). Income resemblance fell substantially across the life course (to 0.38), whereas education resemblance was stable. Psychiatric resemblance changed little across windows. Adjusting psychiatric liability for own SES reduced psychiatric partner resemblance, whereas adjusting SES for own psychiatric status barely affected SES partner resemblance. The reduction in psychiatric partner resemblance had a median of ∼7% before first childbirth but ∼26% across the life course, as diagnosis and SES became increasingly coupled within individuals. What psychiatric partner resemblance represents therefore depends on when it was measured.

## Introduction

Partner resemblance is pervasive across human populations, documented across social, behavioural and health-related traits.^1–6^ It is commonly discussed in terms of assortative mating, the tendency to choose partners with similar traits, but resemblance between established partners need not reflect mate choice. Geographic proximity, shared social environments and overlapping social networks bring similar people together irrespective of preference, and partners may converge, or diverge, in the years after a relationship forms.^2,4^

Which of those processes are involved determines what follows from the resemblance. To the extent that partners resemble each other on heritable traits at the point of union, assortment can generate correlations among trait-increasing alleles, increase resemblance between relatives, and, for a single trait, elevate heritability across generations.^7,8^ Where partners assort on two distinct traits, the same mechanism leads to a genetic correlation between them.^9^ Spouses also transmit a shared set of risk-increasing environmental circumstances alongside their genotypes, so that resemblance can concentrate disease risk in families through both nature and nurture.^10^ Because people tend to partner within similar socioeconomic strata, partner resemblance is also one of the major forces that can reinforce social stratification.^11^

Psychiatric disorders are an informative setting in which to examine these processes. Partner resemblance for disease is generally modest, but psychiatric disorders stand out among clinically relevant traits, showing higher partner correlations than most somatic illnesses.^10,12,13^ Partner correlations for psychiatric disorders are moreover consistent across cultures and persistent across generations, suggesting a general feature of how couples form rather than a local or cohort-specific phenomenon.^14^ Resemblance is considerably stronger for socioeconomic outcomes such as educational attainment and income, which structure the schools, workplaces, and neighbourhoods in which people meet.^2,15^ Within individuals, psychiatric disorders are inversely associated with socioeconomic status, so shared socioeconomic position could generate apparent assortment on psychiatric disorders. Adjusting each partner’s psychiatric liability for their own socioeconomic status has been used to ask how much of the resemblance this accounts for: in Norwegian primary-care records measured 5 to 10 years before a couple’s first child, adjustment for educational attainment removed roughly a third of the partner resemblance in mental health.^13^

What that adjustment removes, however, need not be the same quantity at every point in a couple’s life. The association between psychiatric illness and socioeconomic position runs in both directions: disadvantage can raise the risk of illness, and illness can erode socioeconomic position, so the two become more tightly coupled within individuals as people age.^16,17^ Educational attainment is largely settled before couples form, whereas income continues to change in the years after, so socioeconomic resemblance between partners may itself also shift as earnings trajectories develop. If diagnosis and SES become more tightly linked within individuals over time, adjusting for SES will remove more psychiatric partner resemblance later in life even if partner resemblance itself has not changed. The apparent socioeconomic share can therefore grow without any change in how couples originally formed.

Assortment on socioeconomic status and assortment on psychiatric liability have different consequences for the genetic architecture of psychiatric disorders. If partners sort on psychiatric liability, the alleles associated with that liability become correlated in the next generation. If they sort on socioeconomic status instead, socioeconomic alleles become correlated, and part of that signal will surface as genetic correlation between psychiatric disorders and socioeconomic outcomes, and so as part of the genetic architecture that genome-wide association studies of psychiatric disorders recover.^18,19^ Correlations arising either way can influence estimates derived from such studies.^20^ Which route is operating depends on what partners resembled each other on, and a partner correlation measured after diagnosis and socioeconomic position have become intertwined within individuals cannot distinguish them.

Here we use nationwide Danish register data on 787,658 parental couples to quantify partner resemblance within and across ten psychiatric disorders, educational attainment and income. We estimate resemblance twice, using information accrued up to one year before the birth of the couple’s first child and across the full follow-up, and we adjust in both directions: psychiatric liability for own socioeconomic outcomes, and socioeconomic outcomes for own psychiatric liability. This lets us separate how much psychiatric partner resemblance changes over the life course from how much the socioeconomic share of it changes, to ask whether the overlap between the two domains is symmetric, and to establish whether what psychiatric partner resemblance represents is itself a function of when it was measured.

## Results

We identified 787,658 parental couples from the Danish Civil Registration System, linking individuals who were jointly registered as parents of at least one shared child, from a source population of 5,277,115 individuals born in Denmark between 1969 and 2024. For each partner, we derived ten psychiatric diagnoses, identified from ICD-8 and ICD-10 codes, and two indicators of socioeconomic status (SES), educational attainment and income, over two observation windows: information accrued up to one year before the birth of the couple’s first child (pre-birth), and across the full follow-up period (lifetime).

Within individuals, psychiatric diagnosis was only weakly correlated with SES in the pre-birth window but became substantially more strongly correlated across the life course, particularly for income (Table 1). Among women, having had any psychiatric disorder showed a correlation with their own income of −0.10 pre-birth and −0.42 over the lifetime; the corresponding change for education was smaller (−0.23 to −0.28). Men showed the same pattern, though the lifetime income correlation was weaker (−0.09 to −0.27).

**Table 1:**
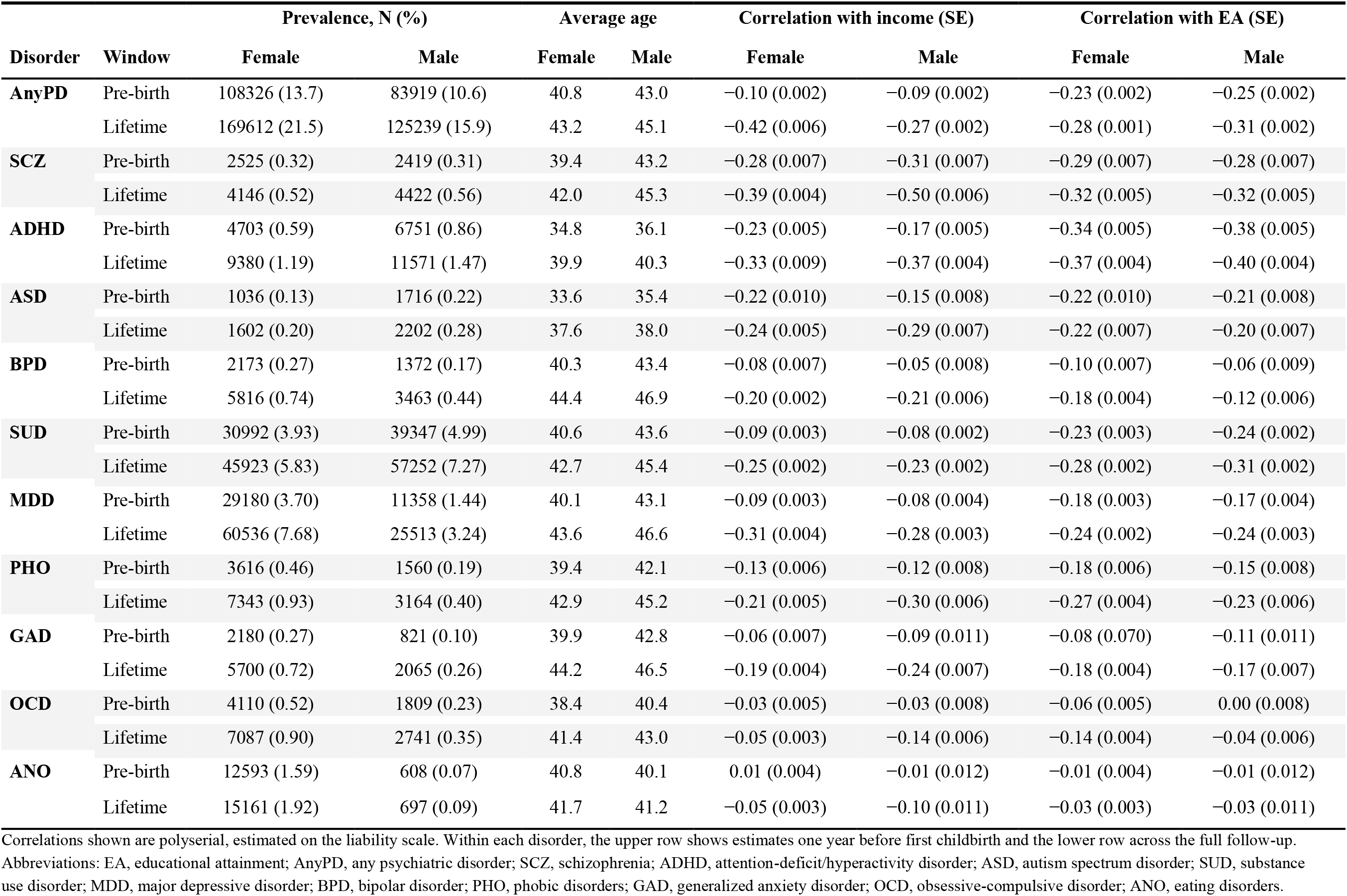
Descriptive statistics of psychiatric disorders and their correlations with income and educational attainment, one year before the birth of the first child (Pre-birth) and across the full follow-up (Life)

| Disorder | Window | Prevalence, N (%) |  | Average age |  | Correlation with income (SE) |  | Correlation with EA (SE) |  |
| --- | --- | --- | --- | --- | --- | --- | --- | --- | --- |
|  |  | Female | Male | Female | Male | Female | Male | Female | Male |
| AnyPD | Pre-birth | 108326 (13.7) | 83919 (10.6) | 40.8 | 43.0 | -0.10 (0.002) | -0.09 (0.002) | -0.23 (0.002) | -0.25 (0.002) |
|  | Lifetime | 169612 (21.5) | 125239 (15.9) | 43.2 | 45.1 | -0.42 (0.006) | -0.27 (0.002) | -0.28 (0.001) | -0.31 (0.002) |
| SCZ | Pre-birth | 2525 (0.32) | 2419 (0.31) | 39.4 | 43.2 | -0.28 (0.007) | -0.31 (0.007) | -0.29 (0.007) | -0.28 (0.007) |
|  | Lifetime | 4146 (0.52) | 4422 (0.56) | 42.0 | 45.3 | -0.39 (0.004) | -0.50 (0.006) | -0.32 (0.005) | -0.32 (0.005) |
| ADHD | Pre-birth | 4703 (0.59) | 6751 (0.86) | 34.8 | 36.1 | -0.23 (0.005) | -0.17 (0.005) | -0.34 (0.005) | -0.38 (0.005) |
|  | Lifetime | 9380 (1.19) | 11571 (1.47) | 39.9 | 40.3 | -0.33 (0.009) | -0.37 (0.004) | -0.37 (0.004) | -0.40 (0.004) |
| ASD | Pre-birth | 1036 (0.13) | 1716 (0.22) | 33.6 | 35.4 | -0.22 (0.010) | -0.15 (0.008) | -0.22 (0.010) | -0.21 (0.008) |
|  | Lifetime | 1602 (0.20) | 2202 (0.28) | 37.6 | 38.0 | -0.24 (0.005) | -0.29 (0.007) | -0.22 (0.007) | -0.20 (0.007) |
| BPD | Pre-birth | 2173 (0.27) | 1372 (0.17) | 40.3 | 43.4 | -0.08 (0.007) | -0.05 (0.008) | -0.10 (0.007) | -0.06 (0.009) |
|  | Lifetime | 5816 (0.74) | 3463 (0.44) | 44.4 | 46.9 | -0.20 (0.002) | -0.21 (0.006) | -0.18 (0.004) | -0.12 (0.006) |
| SUD | Pre-birth | 30992 (3.93) | 39347 (4.99) | 40.6 | 43.6 | -0.09 (0.003) | -0.08 (0.002) | -0.23 (0.003) | -0.24 (0.002) |
|  | Lifetime | 45923 (5.83) | 57252 (7.27) | 42.7 | 45.4 | -0.25 (0.002) | -0.23 (0.002) | -0.28 (0.002) | -0.31 (0.002) |
| MDD | Pre-birth | 29180 (3.70) | 11358 (1.44) | 40.1 | 43.1 | -0.09 (0.003) | -0.08 (0.004) | -0.18 (0.003) | -0.17 (0.004) |
|  | Lifetime | 60536 (7.68) | 25513 (3.24) | 43.6 | 46.6 | -0.31 (0.004) | -0.28 (0.003) | -0.24 (0.002) | -0.24 (0.003) |
| PHO | Pre-birth | 3616 (0.46) | 1560 (0.19) | 39.4 | 42.1 | -0.13 (0.006) | -0.12 (0.008) | -0.18 (0.006) | -0.15 (0.008) |
|  | Lifetime | 7343 (0.93) | 3164 (0.40) | 42.9 | 45.2 | -0.21 (0.005) | -0.30 (0.006) | -0.27 (0.004) | -0.23 (0.006) |
| GAD | Pre-birth | 2180 (0.27) | 821 (0.10) | 39.9 | 42.8 | -0.06 (0.007) | -0.09 (0.011) | -0.08 (0.070) | -0.11 (0.011) |
|  | Lifetime | 5700 (0.72) | 2065 (0.26) | 44.2 | 46.5 | -0.19 (0.004) | -0.24 (0.007) | -0.18 (0.004) | -0.17 (0.007) |
| OCD | Pre-birth | 4110 (0.52) | 1809 (0.23) | 38.4 | 40.4 | -0.03 (0.005) | -0.03 (0.008) | -0.06 (0.005) | 0.00 (0.008) |
|  | Lifetime | 7087 (0.90) | 2741 (0.35) | 41.4 | 43.0 | -0.05 (0.003) | -0.14 (0.006) | -0.14 (0.004) | -0.04 (0.006) |
| ANO | Pre-birth | 12593 (1.59) | 608 (0.07) | 40.8 | 40.1 | 0.01 (0.004) | -0.01 (0.012) | -0.01 (0.004) | -0.01 (0.012) |
|  | Lifetime | 15161 (1.92) | 697 (0.09) | 41.7 | 41.2 | -0.05 (0.003) | -0.10 (0.011) | -0.03 (0.003) | -0.03 (0.011) |

Correlations shown are polyserial, estimated on the liability scale. Within each disorder, the upper row shows estimates one year before first childbirth and the lower row across the full follow-up. Abbreviations: EA, educational attainment; AnyPD, any psychiatric disorder; SCZ, schizophrenia; ADHD, attention-deficit/hyperactivity disorder; ASD, autism spectrum disorder; SUD, substance use disorder; MDD, major depressive disorder; BPD, bipolar disorder; PHO, phobic disorders; GAD, generalized anxiety disorder; OCD, obsessive-compulsive disorder; ANO, eating disorders.

### Partner resemblance before first childbirth

We estimated partner resemblance for psychiatric disorders and SES on the underlying liability scale, accounting for their binary and ordinal measurement scales, respectively (Figure 1). In the pre-birth window, resemblance was high for SES, with correlations of 0.63 for educational attainment and 0.57 for income. Psychiatric disorders showed positive but more moderate correlations, ranging from 0.08 for eating disorders to 0.36 for schizophrenia, with schizophrenia, ADHD, autism spectrum disorder and bipolar disorder showing the strongest resemblance. Having had any psychiatric disorder showed a partner correlation of 0.25.

**Figure 1:**
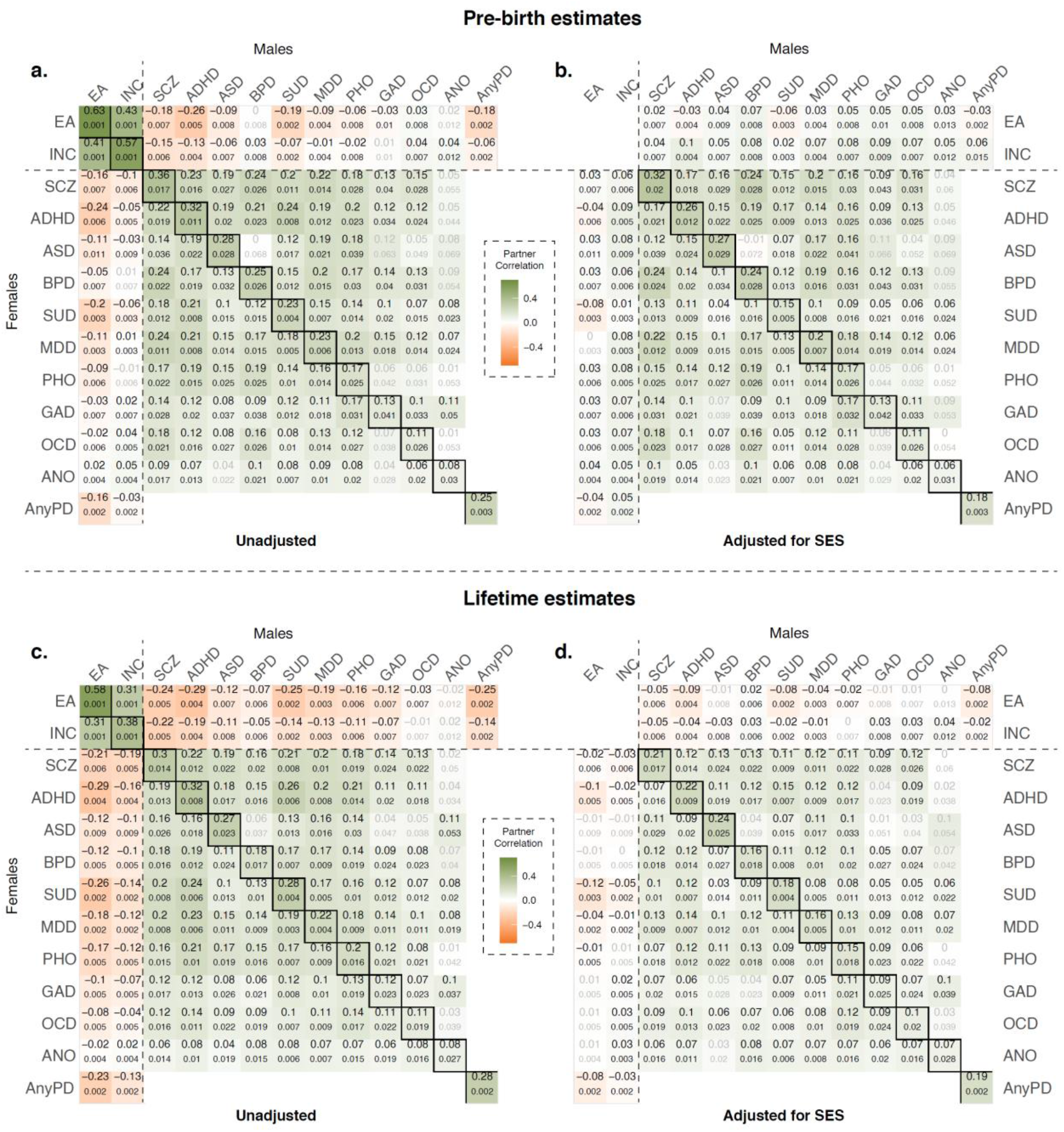
Within- and cross-trait partner correlations for psychiatric disorders and SES, before and after SES adjustment. Rows show estimates measured one year before the birth of the first child (a, b) and across the full follow-up (c, d); columns show partner correlations before (a, c) and after (b, d) adjustment for socioeconomic status (SES). Cells show the correlation between the female partner’s trait (rows) and the male partner’s trait (columns); standard errors are given below each estimate. Diagonal cells show within-trait correlations and off-diagonal cells show cross-trait correlations. SES indicators: EA, educational attainment; INC, income. Psychiatric disorders: SCZ, schizophrenia; ADHD, attention-deficit/hyperactivity disorder; ASD, autism spectrum disorder; BPD, bipolar disorder; SUD, substance use disorder; MDD, major depressive disorder; PHO, phobic disorders; GAD, generalized anxiety disorder; OCD, obsessive-compulsive disorder; ANO, eating disorders; AnyPD, any psychiatric disorder.

Cross-trait correlations between one partner’s educational attainment and the other’s income were also positive (0.41–0.43), whereas cross-partner associations between psychiatric liability and SES were generally negative and stronger for education than for income (Figure 1a). Cross-disorder psychiatric correlations were generally positive but modest, mostly below 0.20 and reaching a maximum of 0.24, for schizophrenia with bipolar disorder and with major depression, and for ADHD with substance use disorder.

Adjusting each partner’s psychiatric liability for their own SES attenuated within-disorder partner correlations to varying degrees (Figure 1b). The correlation for any psychiatric disorder declined from 0.25 to 0.18, a 27% reduction, and within-disorder reductions ranged from approximately 1% to 34%, with the largest for substance use disorder (Extended Data Fig. 1a). Percentage reductions for the least common disorders are computed on small baseline correlations and are correspondingly imprecise, and we do not interpret individual values at the extremes of this range. Cross-disorder correlations were attenuated more strongly than within-disorder correlations, by a median of approximately 20% compared with 7%. Although cross-disorder baseline correlations were generally smaller, this pattern was also evident when considering the absolute change in the correlation coefficients, with median reductions of approximately 0.018 and 0.012 respectively.

Of the two SES indicators, educational attainment contributed more to the attenuation than income. For any psychiatric disorder, the partner correlation was reduced by 29% after adjustment for educational attainment compared with 11% after adjustment for income (Extended Data Figs. 2b and 3b).

Conversely, conditioning SES partner correlations on each partner’s psychiatric status left them essentially unchanged (Extended Data Fig. 4). Adjustment for any psychiatric disorder reduced the partner correlation for educational attainment from 0.63 to 0.62 and for income from 0.570 to 0.566, with similarly small changes after adjustment for individual disorders. The adjustment was therefore strongly asymmetric, and not only in proportional terms: SES adjustment removed about 0.07 of the psychiatric partner correlation, whereas psychiatric adjustment removed about 0.01 of the considerably larger correlation for educational attainment.

### SES accounts for more psychiatric partner resemblance across the life course

Repeating the analysis using lifetime measures showed that psychiatric partner correlations themselves were largely unchanged relative to the pre-birth window (Figure 1c): the correlation for any psychiatric disorder was 0.28, compared with 0.25 pre-birth, and most disorders differed by only a few points. The largest changes were observed for substance use disorder, for which the correlation increased from 0.23 to 0.28, and for schizophrenia and bipolar disorder, for which correlations decreased from 0.36 to 0.30 and from 0.25 to 0.18, respectively (Figs. 1-2).

**Figure 2:**
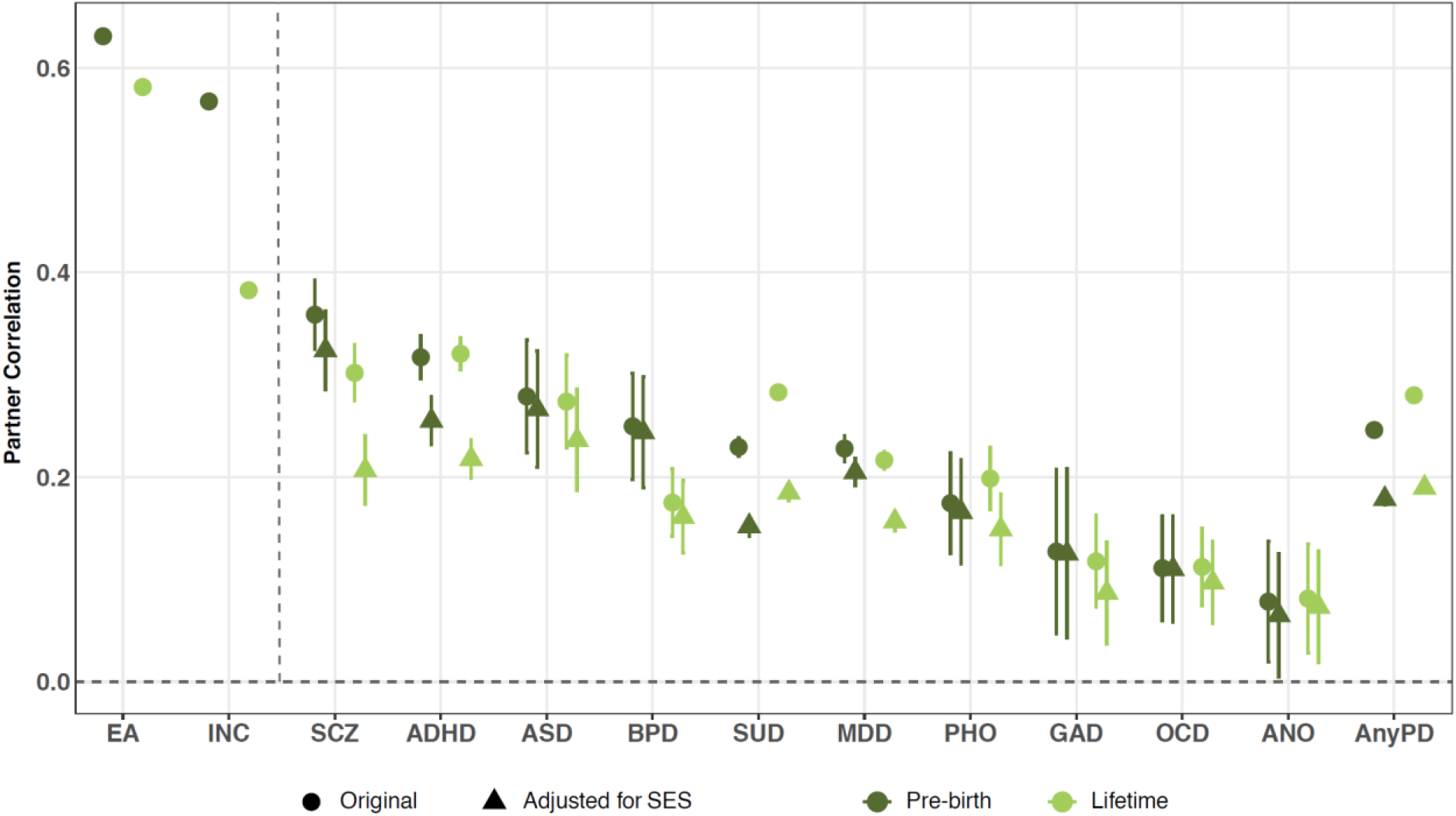
Within-trait partner correlations for psychiatric disorders and SES across observation windows, before and after SES adjustment. Points show unadjusted correlations and correlations after adjusting each partner’s psychiatric liability for their own SES. Estimates are shown for information accrued up to one year before first shared childbirth and across the full available follow-up. Error bars indicate 95% confidence intervals. SES indicators: EA, educational attainment; INC, income. Psychiatric disorders: SCZ, schizophrenia; ADHD, attention-deficit/hyperactivity disorder; ASD, autism spectrum disorder; BPD, bipolar disorder; SUD, substance use disorder; MDD, major depressive disorder; PHO, phobic disorders; GAD, generalized anxiety disorder; OCD, obsessive-compulsive disorder; ANO, eating disorders; AnyPD, any psychiatric disorder.

SES adjustment produced substantially greater attenuation of lifetime than pre-birth correlations (Figure 1d). Within-disorder reductions ranged from approximately 8% to 35% (Extended Data Fig. 1b), with a median of approximately 26% compared with 7% pre-birth. The increase was clearest for schizophrenia, where attenuation rose from 10% to 32%, and major depression, from 10% to 28%. Educational attainment again accounted for more of the attenuation than income, reducing the lifetime correlation for any psychiatric disorder by 31%, compared with 13% for income (Extended Data Figs. 2d and 3d). Cross-partner associations between SES and psychiatric liability were generally stronger in the lifetime window, but the proportional reductions produced by SES adjustment were larger before childbirth (Extended Data Figs. 1–3)

SES partner resemblance also differed across measurement windows. Income resemblance was substantially lower using lifetime measures than pre-birth (0.38 versus 0.57), whereas educational attainment, which is largely established earlier in life, was comparatively stable (0.58 versus 0.63). The lower lifetime income correlation may reflect divergence in partners’ earnings after childbirth rather than differences in partner selection. Conditioning SES on psychiatric status again left partner correlations largely intact, though the reductions were somewhat larger than pre-birth: adjustment for any psychiatric disorder lowered the partner correlation for educational attainment from 0.58 to 0.56 and for income from 0.38 to 0.37 (Extended Data Fig. 4). The asymmetry therefore persists across windows, while the reverse adjustment increases in parallel with the primary one, consistent with diagnosis and socioeconomic status becoming more closely coupled within individuals over time.

## Discussion

In 787,658 Danish parental couples we observed substantial partner resemblance for psychiatric disorders, ranging from 0.08 for eating disorders to 0.36 for schizophrenia, against a background of considerably stronger resemblance for educational attainment (0.63) and income (0.57). These estimates are consistent with earlier evidence of non-random mating within and across psychiatric disorders,^12–14^ and extend it to a design that contrasts information accrued before couples had their first child with information accumulated across the full follow-up. Three findings structure our interpretation. First, measured socioeconomic status captured an appreciable but minority share of psychiatric partner resemblance in the pre-birth window. Second, this overlap was strongly asymmetric: socioeconomic status accounted for a meaningful share of psychiatric partner resemblance, whereas psychiatric status accounted for only a small fraction of the considerably stronger partner resemblance in socioeconomic status. Third, the share of psychiatric partner resemblance captured by socioeconomic status grew substantially when traits were measured across the life course, even though the psychiatric partner correlations themselves changed little.

In the pre-birth window, adjusting each partner’s psychiatric liability for their own education and income reduced the partner correlation for any psychiatric disorder from 0.25 to 0.18, a 27% attenuation, with within-disorder reductions ranging from approximately 1% to 34% and a median of about 7%. Education accounted for more of this than income (29% versus 11%), consistent with education being largely completed before couples form and with its role in structuring the schools, workplaces and neighborhoods in which partners meet.^11^ Conditioning in the opposite direction left SES resemblance largely intact, reducing the correlation for educational attainment from 0.63 to 0.62. The covariance structure is therefore more compatible with psychiatric resemblance being partly embedded within a broad socioeconomic axis of partner sorting than with SES resemblance arising as a consequence of psychiatric assortment. Most within-disorder resemblance nonetheless persisted after adjustment, so education and income alone cannot account for it; assortment on psychiatric characteristics or correlated behavioural traits, geographic and social homogamy, and overlapping social networks are all likely to contribute. Analyses using sibling and in-law data indicate that partner resemblance across a range of traits deviates from direct assortment on the observed phenotype,^13^ so the residual resemblance we observe should not be read as direct assortment on diagnosis. Education and income are, in any case, only partial indicators of socioeconomic position, so the share they remove is a lower bound on the overlap, and the share they leave behind should not be read as independent of socioeconomic position.

Across the two measurement windows, psychiatric partner correlations were largely stable, with any psychiatric disorder moving from 0.25 to 0.28 and most disorders differing by only a few points. Within individuals, however, psychiatric status became more strongly coupled to socioeconomic position, particularly income: the correlation between any psychiatric disorder and own income shifted from −0.10 to −0.42 in women and from −0.09 to −0.27 in men, whereas the changes for education were small (−0.23 to −0.28 and −0.25 to −0.31). Correspondingly, SES adjustment removed considerably more of the lifetime correlations, with the median within-disorder reduction rising from about 7% to 26% and the clearest increases for schizophrenia (10% to 32%) and major depression (10% to 28%). The degree of psychiatric partner resemblance thus changed relatively little, while the share of it captured by each partner’s socioeconomic position became substantially greater.

The larger attenuation in the lifetime window needs careful interpretation. Adjustment for SES is not a decomposition of psychiatric partner resemblance into socioeconomic and non-socioeconomic components: its effect depends on how strongly psychiatric status and SES are associated within each partner, as well as on their associations across partners. These relationships differ substantially between our measurement windows. In particular, psychiatric status became much more strongly associated with socioeconomic position within individuals across the life course, while unadjusted psychiatric partner correlations changed little. The greater attenuation after SES adjustment in the lifetime window therefore does not imply that socioeconomic assortment played a greater role in generating the original partner resemblance. Instead, it may partly reflect the stronger association between psychiatric status and socioeconomic position later in life. Alternatively, lifetime measures ascertain more cases and may capture the same underlying liability more completely.

Income illustrates the same broader principle. Partner resemblance fell from 0.57 before childbirth to 0.38 across follow-up, whereas education was comparatively stable (0.63 to 0.58). This is unlikely to mean that couples somehow “un-assort” on income; rather, earnings trajectories may diverge after childbirth, and our use of maximum observed income may accentuate this divergence, since women’s earnings more often plateau after a first child while men’s continue to rise. Observed partner correlations are therefore not timeless indices of mate choice: they reflect the traits as measured at a particular stage of life and may combine assortment when couples formed, social homogamy, convergence after partnering, and later consequences of the traits themselves.

A population-wide study of Norwegian first-time parents measured primary-care diagnoses 10 to 5 years before the birth of the first child, and found that partner correlations in mental health rose substantially when the same couples were assessed later as established partners.^13^ Our pre-birth estimates resemble their later rather than their earlier window, consistent with the increase they document having largely occurred before our first observation. Read together, the two designs suggest that psychiatric partner resemblance builds during the years around union formation and is largely in place by the time couples have a first child. Our partner correlations are generally higher than theirs, consistent with specialist psychiatric registers capturing a more severe spectrum than primary-care records; the exception is substance use disorder, where their drug-specific definition excludes the alcohol-related diagnoses included in our category.

Estimates for several disorders have not previously been available at this scale: a meta-analysis of partner correlations across 22 traits could not include schizophrenia or bipolar disorder because the available studies yielded underpowered contingency tables.^2^ Where that meta-analysis does provide benchmarks, our estimates are broadly consistent, with partner correlations of 0.23 for major depression against a meta-analytic 0.15 and 0.13 for generalized anxiety against 0.17.

Within our own estimates, the few disorders whose partner correlations changed considerably across windows moved in opposite directions. The increase for substance use disorder (0.23 to 0.28) is compatible with convergence through shared partner environments,^21^ while the decreases for schizophrenia (0.36 to 0.30) and bipolar disorder (0.25 to 0.18) indicate that cases diagnosed later are less concordant between partners, plausibly reflecting earlier age of onset and a greater contribution of cases already expressed around the time couples formed.^22^

A more general distinction runs across disorders. Cross-disorder correlations attenuated more strongly under SES adjustment than same-disorder correlations, by a median of about 20% compared with 7%, a difference that holds on the absolute scale as well. This complements a recent Danish population-wide analysis, in which much of the cross-disorder resemblance was attributable to psychiatric comorbidity.^10^ The same contrast has been reported in Norwegian primary-care data, where adjustment for education reduced cross-trait mental-health correlations by 41% against 31% for within-trait correlations.^13^ Partners sharing the same diagnosis may therefore reflect mechanisms relatively specific to that phenotype, whereas the broader tendency for one partner’s disorder to predict a different disorder in the other may more strongly reflect nonspecific social stratification or correlated traits.

Because both psychiatric disorders and socioeconomic outcomes are heritable, the mating patterns documented here have implications beyond their phenotypic description. A recent Danish population-wide study using the same registers found that observed partner similarity was associated with higher psychiatric prevalence and family-based heritability than under simulated random mating.^10^ The present results refine the interpretation of such effects by showing that the underlying partner resemblance is partly embedded in socioeconomic sorting and depends on the measurement window. To the extent that partner resemblance reflects assortment on heritable liability, it is expected to generate correlations among trait-increasing alleles, alter resemblance among relatives, and, in the cross-trait case, contribute to genetic covariance between socioeconomic and psychiatric traits in subsequent generations.^9,20^ Our results do not quantify that contribution, and we make no claim about how much of any observed genetic correlation is mating-induced. They do, however, indicate that the phenotypic quantity such models take as input is not window-invariant: partner resemblance for income and the psychiatric–socioeconomic cross-terms both differ substantially between our two windows, even though psychiatric within-trait resemblance does not. The choice of measurement window therefore determines whether such models describe sorting at the time couples form or the accumulated association between traits later in life.

Several limitations should be kept in mind when interpreting our results. Our sample comprises parental couples, so estimates condition on shared fertility — itself downstream of both partners’ traits — which may matter most for disorders associated with reduced fertility such as schizophrenia. The pre-birth window postdates the start of most relationships, by a median of roughly four years in comparable Norwegian data,^13^ so our estimates describe established rather than newly formed couples, and it is anchored to age at first birth, which varies across disorders. Outpatient contacts are recorded only from 1995, so ascertainment in this window varies across cohorts. Lifetime measures span each individual’s full follow-up, including any period after the union dissolved. Individuals with children by more than one partner contribute to multiple couples, and standard errors do not account for this clustering. Register diagnoses capture treated disorder rather than liability, and because help-seeking is a behaviour partners may share, some resemblance in recorded diagnosis need not reflect resemblance in liability. Income was defined as a running maximum, which may accentuate post-childbirth divergence between partners. Adjustment for own SES is descriptive rather than causal, and is complicated by SES being partly downstream of illness. Finally, the source population excludes immigrants, Denmark’s compressed income distribution and universal healthcare limit generalization, and results pertain to opposite-sex parental couples only.

Psychiatric partner resemblance is substantial, largely in place by the time couples have their first child, and partly embedded within considerably stronger socioeconomic sorting. Its apparent socioeconomic component, however, depends on when the constituent traits are measured, because psychiatric illness and socioeconomic position become increasingly intertwined within individuals over the life course. Partner correlations, and especially what statistical adjustment appears to explain, should be interpreted accordingly.

## Methods

Using Danish nationwide register data linked at the individual level through the unique personal identification number assigned to all residents, we conducted a population-based retrospective cohort study. The Danish Civil Registration System (CRS),^23^ established in 1968, provided the demographic backbone for the study, including information on vital status, migration, residence, and parental and marital links. We used the CRS to define the source population as the 5,277,115 individuals born in Denmark between January 1, 1969, and 2024. Parental couples were identified through registered parent–child links as pairs of individuals who had at least one child together, resulting in 787,658 parental couples; because both partners must have reached parenthood within the observation period, these couples are drawn from the earlier part of the source-population birth range. Individuals who died or emigrated were retained in the study, with all information recorded up to their last available observation included in the respective observation windows. At the end of the pre-birth observation window, 1 year before the birth of the couple’s first child, the median age was 28.8 years (IQR, 25.8–32.2) among fathers and 27.1 years (IQR, 24.0–30.2) among mothers. At the end of available follow-up, the corresponding median ages were 44.6 years (IQR, 37.9–50.3) and 42.1 years (IQR, 35.8–48.2), respectively.

Psychiatric diagnoses were obtained from the Danish National Patient Register (NPR)^24^ and the Danish Psychiatric Central Research Register (PCRR)^25^, which together provide nationwide information on inpatient psychiatric contacts and, from 1995, outpatient and emergency contacts. These registers enabled identification of psychiatric disorders coded according to ICD-8 and ICD-10. Psychiatric outcomes were defined for ten disorders: schizophrenia (SCZ), attention-deficit/hyperactivity disorder (ADHD), autism spectrum disorder (ASD), substance use disorder (SUD), major depressive disorder (MDD), phobic disorders (PHO), bipolar disorder (BPD), eating disorders (ANO), generalized anxiety disorder (GAD), and obsessive-compulsive disorder (OCD), together with a composite measure of any psychiatric disorder (AnyPD). ADHD was defined by ICD-10 code F90.0 only. Each disorder was coded as a binary indicator of whether the individual had received the diagnosis; corresponding ICD codes are provided in Table 2.

**Table 2.** ICD Code List for Psychiatric Disorders.

| **Disorder** | **ICD 10** | **ICD 8** |
| --- | --- | --- |
| **SUD** | F10-F19 | 291, 294.39, 303, 304 |
| **SCZ** | F20 | 295 |
| **MDD** | F32, F33 | 296.09, 296.29, 298.09, 300.49 |
| **OCD** | F42 | 300.39 |
| **PHO** | F40.0, F40.1, F40.2 |  |
| **GAD** | F41.1 |  |
| **ANO** | F50 | 305.60, 306.50 , 306.58 , 306.59 |
| **ASD** | F84.0, F84.1, F84.5, F84.8, F84.9 |  |
| **ADHD** | F90.0 |  |
| **BPD** | F30, F31 | 296.19, 296.39, 298.19 |
| **AnyPD** | F00–F99 | 290–315 |

Note: ICD codes represent broad categories and include all sub-diagnoses.

Socioeconomic status was measured using educational attainment and income. Educational attainment was obtained from the Danish Education Registers, which provide regularly updated nationwide, individual-level information on enrolment, completed education, and educational trajectories.^26^ It was defined as the highest completed level of education and harmonized into eight ordinal categories based on the International Standard Classification of Education 2011 (ISCED-2011). Income was derived from Danish income registers compiled by Statistics Denmark and defined as the maximum annual taxable income observed up to the end of the relevant observation window.^27^ We treated income as an ordinal variable and categorized it into ten sex-specific deciles to account for sex differences in income distributions.

All psychiatric and socioeconomic measures were defined over two observation windows. The pre-birth window comprised all information accrued up to one year before the birth of the couple’s first child: psychiatric diagnoses recorded up to that point, the maximum annual taxable income observed up to that point, and the highest completed education recorded up to that point. Lifetime measures used all information available across the full observation period, including psychiatric diagnoses recorded at any time during follow-up, maximum observed income, and highest completed education. The lifetime window extended the pre-birth window by incorporating information recorded subsequently.

### Quantifying Partner Resemblance

We estimated partner resemblance under a liability threshold framework using structural equation modeling (SEM), implemented in the lavaan package in R.^28^ This approach accommodates binary and ordinal traits and allows estimation of partner correlations on the underlying latent liability scale.^20^

### Baseline Estimation of Partner Resemblance

Psychiatric diagnoses were modeled as binary variables, assuming that observed case status reflects crossing a threshold on an underlying normally distributed liability. The same liability-threshold framework was applied to the ordinal SES traits, educational attainment and income. For binary outcomes, the liability model assumes a single threshold:

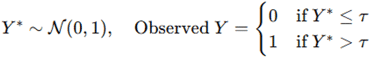

For ordinal variables, the same model is extended to include multiple thresholds separating adjacent categories. In the baseline model, for psychiatric disorders, birth year and sex were included as covariates to account for cohort and sex differences in follow-up time, diagnostic practices, healthcare access, and case detection. For SES traits, education and income were adjusted for age to account for demographic and period-related shifts in educational attainment and earnings.

### Adjustment Models

To examine the extent to which psychiatric partner resemblance was statistically accounted for by SES, and vice versa, we extended the baseline liability-threshold SEM by introducing reciprocal adjustment between psychiatric and SES traits.

For psychiatric partner resemblance adjusted for SES, each partner’s psychiatric liability was regressed on their own educational attainment and income. The residual partner correlation therefore reflects the resemblance remaining after removing the component of each partner’s liability that is linearly predicted by their own SES; it is a descriptive residual and does not by itself identify assortment on psychiatric liability. We repeated this procedure adjusting for education and income separately, to estimate the contribution of each SES indicator.

For SES partner resemblance adjusted for psychiatric status, each partner’s educational attainment or income was regressed on their own psychiatric status within the same SEM framework, and the residual partner correlation represents SES resemblance after accounting for psychiatric status.

In both directions, adjustment was applied to all cells of the correlation matrix, so residual estimates are reported for within-trait, cross-disorder and cross-partner SES–psychiatric associations alike.

## Data Availability

The individual-level Danish register data used in this study cannot be made publicly available owing to data protection regulations. Researchers may apply for access to the underlying register data through the relevant Danish authorities, including Statistics Denmark and the Danish Health Data Authority, subject to the required approvals and data-use agreements.

## Acknowledgements

This research was supported by the US National Institutes of Health study on extreme major depressive disorder (grant No. R01 MH123724) and by the European Union’s Horizon 2020 research and innovation programme under grant agreements No. 847776 and No. 964874. A.A. is supported by The Amsterdam UMC Fellowship.

---

**Extended Data Fig. 1:**
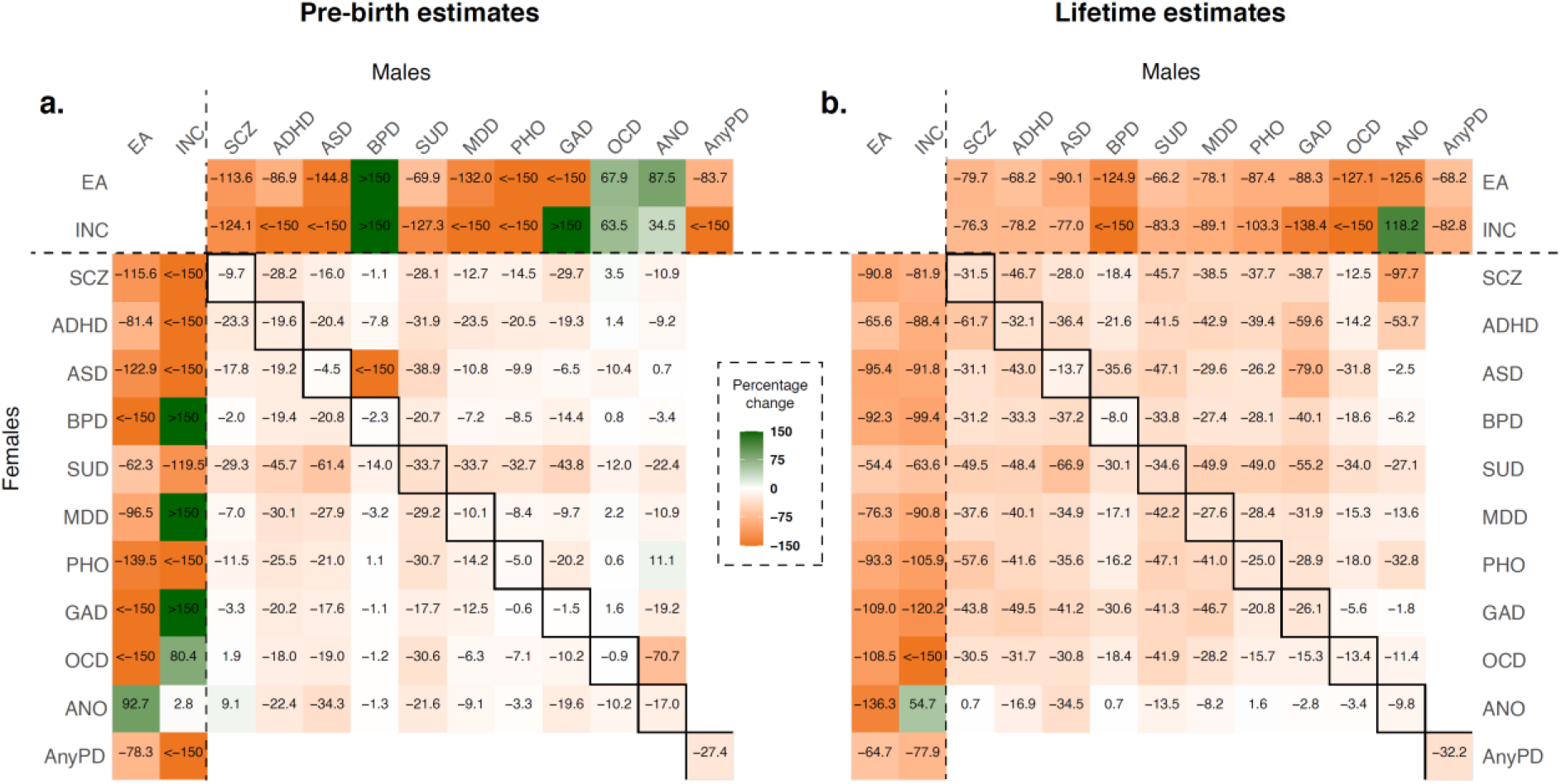
Percentage change in spousal correlations after adjustment for socioeconomic status. Percentage change in spousal correlations after adjusting each partner’s psychiatric traits for their own educational attainment and income, for measures obtained one year before the birth of the first child (**a**) and across the full follow-up (**b**). Rows correspond to female traits and columns to male traits. Values show percentage change relative to the corresponding unadjusted spousal correlation; values exceeding ±150% are displayed as >150 or <−150. SES indicators: EA, educational attainment; INC, income. Psychiatric disorders: SCZ, schizophrenia; ADHD, attention-deficit/hyperactivity disorder; ASD, autism spectrum disorder; BPD, bipolar disorder; SUD, substance use disorder; MDD, major depressive disorder; PHO, phobic disorders; GAD, generalized anxiety disorder; OCD, obsessive-compulsive disorder; ANO, eating disorders; AnyPD, any psychiatric disorder.

**Extended Data Fig. 2:**
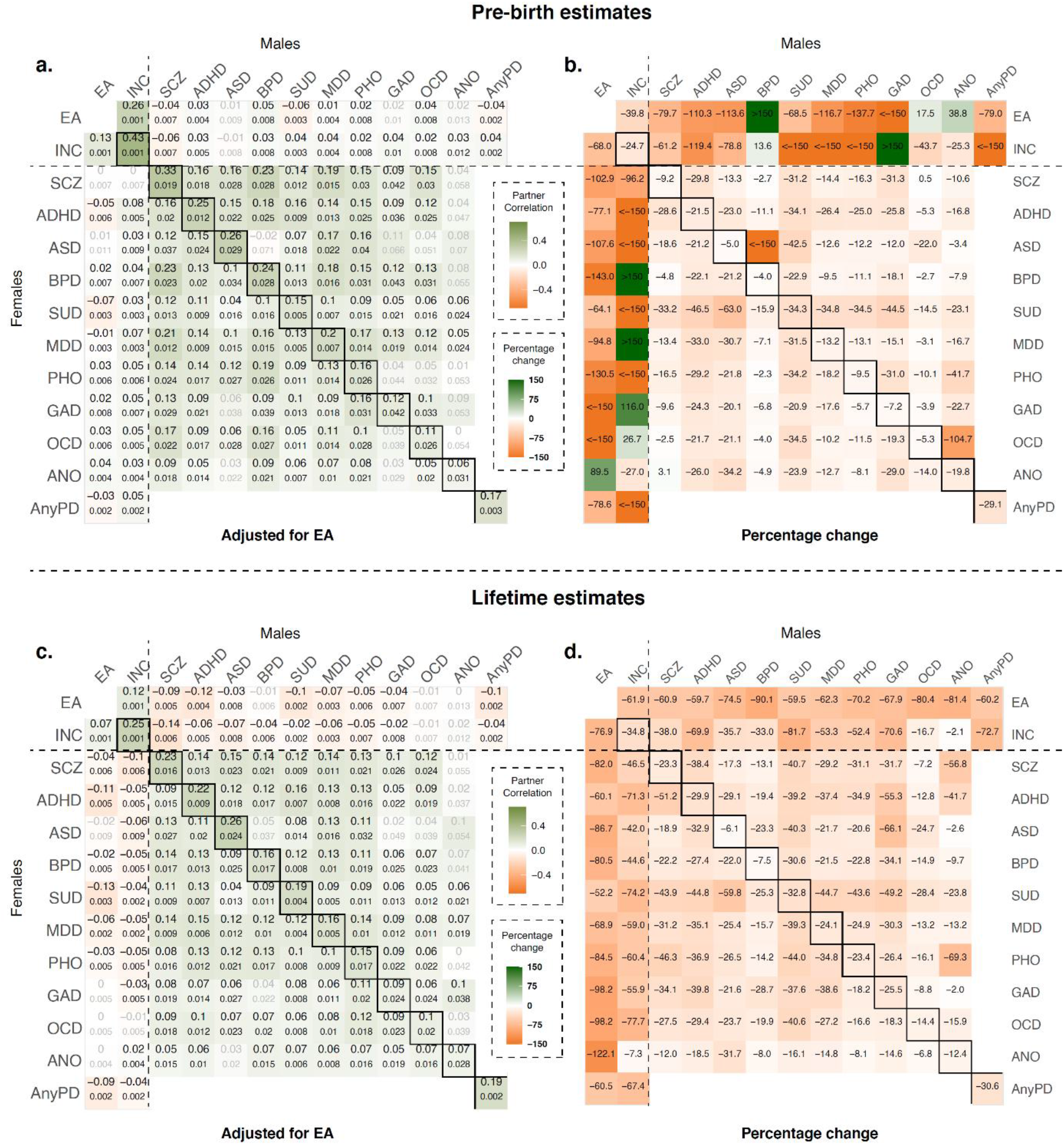
Spousal correlations after adjustment for educational attainment. Spousal correlations after adjusting each partner’s psychiatric traits for their own educational attainment (**a,c**) and the corresponding percentage change relative to the unadjusted correlations (**b,d**), for measures obtained one year before the birth of the first child (**a,b**) and across the full follow-up (**c,d**). Rows correspond to female traits and columns to male traits. Numbers below correlation estimates indicate standard errors. Percentage changes exceeding ±150% are displayed as >150 or <−150. SES indicators: EA, educational attainment; INC, income. Psychiatric disorders: SCZ, schizophrenia; ADHD, attention-deficit/hyperactivity disorder; ASD, autism spectrum disorder; BPD, bipolar disorder; SUD, substance use disorder; MDD, major depressive disorder; PHO, phobic disorders; GAD, generalized anxiety disorder; OCD, obsessive-compulsive disorder; ANO, eating disorders; AnyPD, any psychiatric disorder.

**Extended Data Fig. 3:**
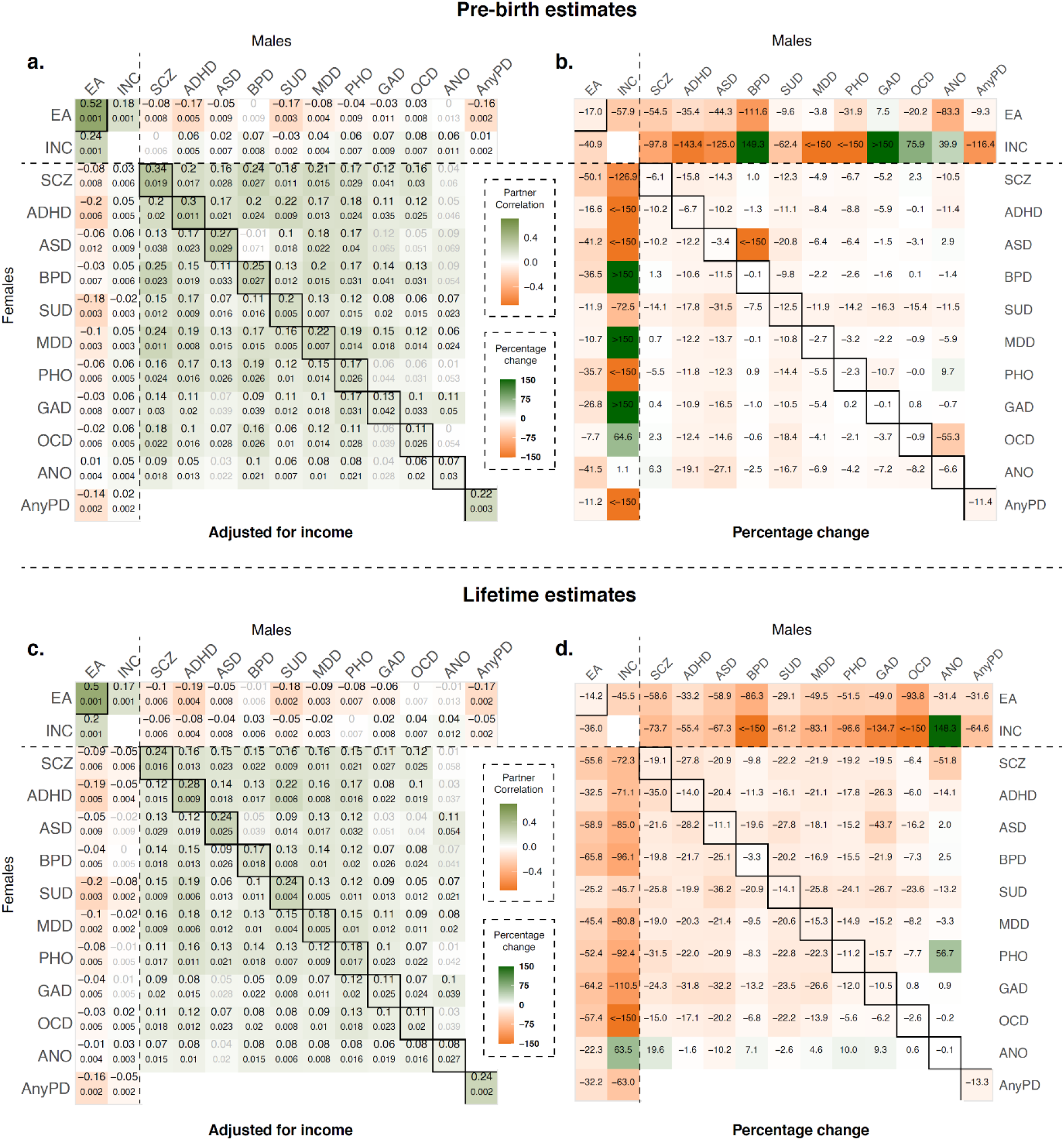
Spousal correlations after adjustment for income. Spousal correlations after adjusting each partner’s psychiatric traits for their own income (**a,c**) and the corresponding percentage change relative to the unadjusted correlations (**b,d**), for measures obtained one year before the birth of the first child (**a,b**) and across the full follow-up (**c,d**). Rows correspond to female traits and columns to male traits. Numbers below correlation estimates indicate standard errors. Percentage changes exceeding ±150% are displayed as >150 or <−150. SES indicators: EA, educational attainment; INC, income. Psychiatric disorders: SCZ, schizophrenia; ADHD, attention-deficit/hyperactivity disorder; ASD, autism spectrum disorder; BPD, bipolar disorder; SUD, substance use disorder; MDD, major depressive disorder; PHO, phobic disorders; GAD, generalized anxiety disorder; OCD, obsessive-compulsive disorder; ANO, eating disorders; AnyPD, any psychiatric disorder..

**Extended Data Fig. 4:**
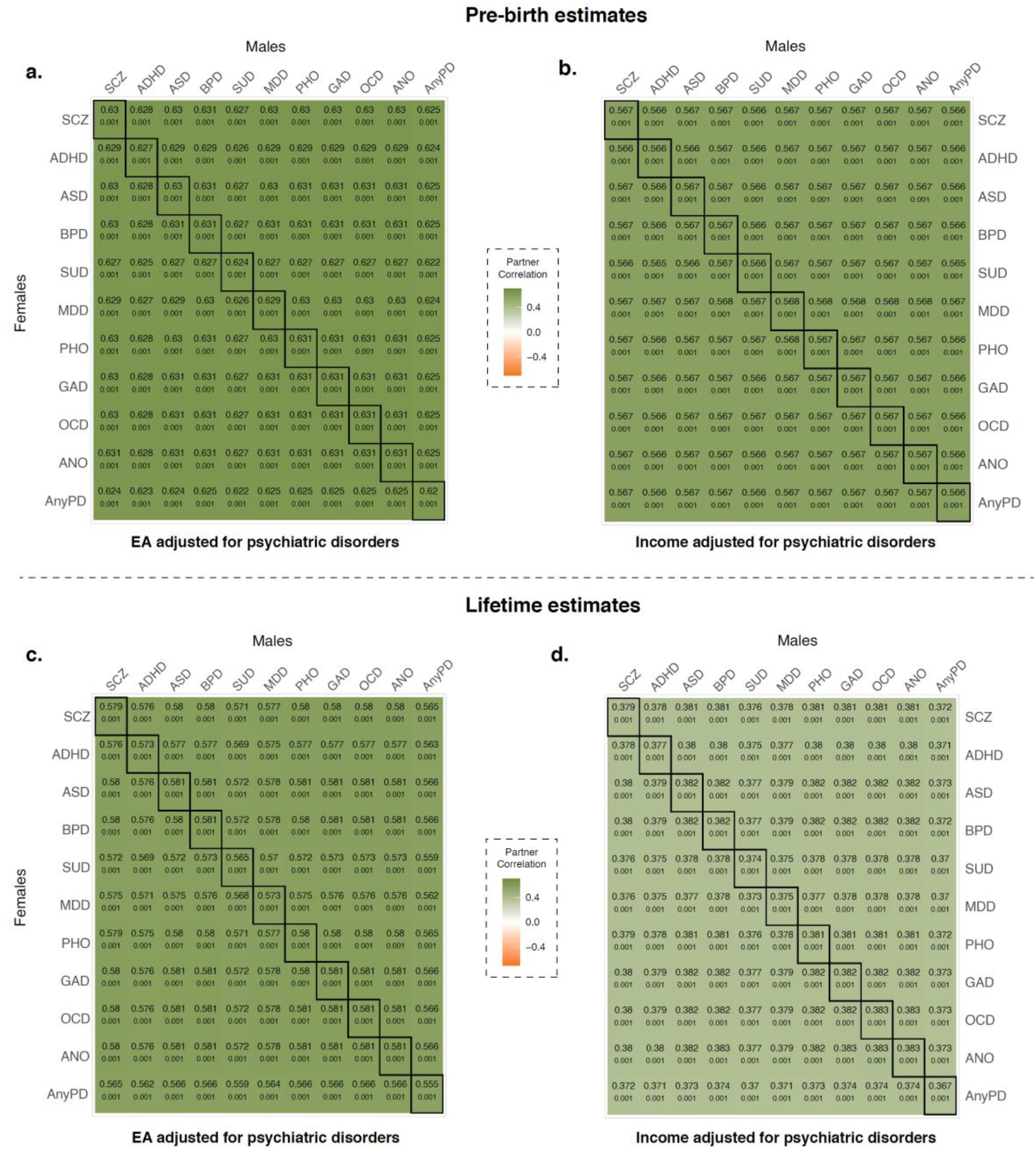
Partner correlations for socioeconomic status after adjustment for psychiatric status. Partner correlations for educational attainment (**a,c**) and income (**b,d**) after regressing each partner’s socioeconomic outcome on their own psychiatric status, for measures obtained one year before the birth of the first child (**a,b**) and across the full follow-up (**c,d**). Rows correspond to the disorder used to adjust the female partner’s socioeconomic outcome and columns to the disorder used to adjust the male partner’s; boxed diagonal cells therefore show adjustment for the same disorder in both partners. Numbers below correlation estimates indicate standard errors. Unadjusted partner correlations were 0.63 (educational attainment) and 0.57 (income) pre-birth, and 0.58 and 0.38 across the full follow-up. EA, educational attainment; SCZ, schizophrenia; ADHD, attention-deficit/hyperactivity disorder; ASD, autism spectrum disorder; BPD, bipolar disorder; SUD, substance use disorder; MDD, major depressive disorder; PHO, phobic disorders; GAD, generalized anxiety disorder; OCD, obsessive-compulsive disorder; ANO, eating disorders; AnyPD, any psychiatric disorder.

